# Influence of socioeconomic, geographic, climatic, and infrastructure factors with dengue fever in Colombia from 2015 to 2020: A Spatial Generalized Additive Mixed Model

**DOI:** 10.64898/2026.01.15.26344190

**Authors:** Sergio Moreno-López, Lucia C. Pérez-Herrera, Olga L. Sarmiento, Luis Fernando Gutierrez, Iveth Mier, Augusto Peñaranda

## Abstract

**Background:** Dengue virus infection remains among the most prevalent infectious diseases globally, with an estimate of 100–400 million cases occurring each year. Prior ecological studies have documented spatial and temporal correlations between dengue occurrence and either socioeconomic or climatic conditions independently. The aim of this study was to analyze the association between multidimensional typologies, including socioeconomic, geographic, climatic, and infrastructure factors and the rate of dengue cases in Colombia.

**Methods:** An analytical, observational, ecological study with repeated measures that included six years of aggregated quantitative data study from 2015 to 2020, based on national registry data from several sources. A spatial Generalized Additive Mixed Model (GAMM) was used to identify the probable factors associated with the frequency of dengue in Colombia. Socioeconomic, infrastructure, geographic, and climatic factors included Multidimensional Poverty Index (MPI), altitude, population density, road density, education and public services coverage, altitude, temperature, precipitation, relative humidity, and presence of El Niño-Southern Oscillation (ENSO) in the period of analysis.

**Results:** During the study period, 448,774 cases of dengue fever were reported in Colombia, with overall monthly median of 6280 cases (IQR: 2842-9278). The highest number of cases was reported in 2019 with 118,956 cases. Higher levels of drinkable water service coverage, education access coverage, and relative humidity were negatively associated with dengue frequency, showing lower case frequency. While higher GDP per capita and greater participation in the subsidized health system were associated with higher incidence of dengue, although the latter showed substantial variability in extreme values. Precipitation and maximum and minimum temperatures showed unimodal associations, with risk concentrated at intermediate values, while altitude and population density revealed complex multimodal patterns. Municipal performance, a composite index that assesses the efficiency and management capacity of local governments, showed a negative association with dengue occurrence, indicating lower risk in better-performing municipalities.

**Conclusions:** A complex interaction was found between socioeconomic, geographic, climatic, and infrastructure factors. These findings may highlight the need for multidimensional approaches in dengue prevention and control policies, that consider the specific conditions of each community. Research is required to better understand the factors that affect the dynamics of dengue. Factors identified at this local level may be useful in different geographical regions, especially in the context of climate change related vulnerabilities.

**Author summary:** Dengue fever affects millions worldwide, with Colombia reporting over 150,000 cases in 2023. Understanding what drives dengue transmission is crucial for prevention, but most studies focus on either climate or social factors alone. We studied how multiple factors such as poverty, access to services, climate conditions, and government capacity relate to dengue patterns across Colombian municipalities from 2015-2020. We found that dengue occurrence is associated with a complex mix of factors. Warmer temperatures and moderate rainfall were associated with more cases, but so were factors like limited access to clean water and education. Surprisingly, areas with higher economic development also reported more cases, possibly explained by better disease detection and diagnosis due to better healthcare access. Well-performing local governments were associated with lower disease frequency. These findings suggest that fighting dengue requires comprehensive approaches that go beyond mosquito control. Communities need improved water access, better education, and strong local governance. As climate change progresses, understanding these multiple vulnerabilities becomes even more important to protect vulnerable populations, particularly in tropical regions where dengue is endemic.

## Introduction

The World Health Organization (WHO) estimates 100-400 million dengue virus infections occurring each year, with a mortality rate of 2.5% [1]. Currently, the medical treatment for dengue is based on symptomatic management, but timely detection and access to medical care can potentially decrease mortality rates to less than 1% [1,2]. By the first half of 2020, more than 1.6 million of dengue cases had been reported in Latin America, with higher frequency rates in Brazil (1,040,481 cases); Paraguay (218,798 cases); Bolivia (82,460 cases); and Argentina (79,775 cases) [2]. In 2023, Colombia reported more than 150,000 cases, and an increase in the incidence of dengue fever, with 387,2 cases per 100,000 inhabitants [3]. Likewise, recent studies have highlighted the increase in the incidence of this disease worldwide, despite calls for vector management, prevention, and control [4]. By the end of 2016, a total of 291,964 cases of dengue associated with outbreaks had been reported, of which 72.4% of dengue patients were reported in the Western Pacific region, followed by the Americas region (19.4%), the Southeast Asia region (4.8%), the Eastern Mediterranean region (1.5%), the European region (1.5%), and the African region (0,3%) [5].

In the last decades, the increase in dengue incidence and the unavailability of treatments for advanced stages have increased its global burden and represent a significant economic impact on affected countries. This scenario is particularly important in regions of the world that are most vulnerable to climate change, such as Asia, Africa, and Latin America, which share socioeconomic characteristics and climatic factors [6,7]. Regarding the average annual costs of dengue in Latin America, this amount is estimated to be $3 billion in both direct and indirect costs and losses in Disability-Adjusted Life Years (DALYs) [8,9]. On the other hand, the reporting and surveillance of dengue cases globally is limited in regions such as sub-Saharan Africa or the Eastern Mediterranean [1,10]; regions where an increase in the dissemination of *Aedes* mosquitoes is expected in the next 50 years [11,12].

Moreover, the increase in the global incidence of dengue has been attributed, in part, to infrastructural factors related to social inequality such as the lack of access to public services and the decrease in the implementation of preventive health programs [13,14], as well as high levels of poverty [15,16], governmental deficiencies, and low institutional effectiveness [6,17]. Overall, this scenario is also exacerbated by the climatic variability mainly attributed to climate change [11,18,19]. Current literature discusses the complex interactions among socioeconomic, geographical, climatic factors that may influence both the biology, reproductive dynamics, and survival of the vector, as well as the infrastructure-related and intermediate social determinants of the populations that are most vulnerable to this disease [20,21]. It is crucial to identify how the combined effect of multiple socioeconomic, climatic, and governmental factors affect the dynamics of dengue, as the interaction among them may exacerbate the frequency and widespread patterns of the disease in many territories worldwide [22,23]. Therefore, this study aims to assess the possible impact of socioeconomic, geographic, climatic, and infrastructure factors on the frequency of dengue, all within a framework of vulnerabilities present in the Colombian territory.

## Materials and methods Ethics Statement

This study was approved by the Ethics Committee of the Hospital Universitario Fundación Santa FE (CCEI-15374–2023).

### Study design

Colombia has a population of 52.6 million inhabitants according to the 2020 national estimate from the National Administrative Department of Statistics (DANE). The country is divided into 32 departments, which are first-level territorial entities, and a total of 1,099 municipalities, which are second-level administrative divisions within departments. Each municipality has local governance structures such as a mayor and municipal council. Dengue surveillance and public health control measures are implemented at both the national (Ministry of Health) and local (municipal health secretariats) levels. Local governments collaborate with both the National Health Institute (INS) and regional health institutions to monitor diseases like dengue [24,25]. In terms of altitude, most of Colombian territory ranges between 1,000 and 2,000 meters above sea level, and its average temperatures range from 11 to 17°C. [26]. In terms of human development, Colombia ranks in the 88th position worldwide and had a Human Development Index (HDI) of 0.756 in 2020, categorizing it as having a middle to high level of human development. However, the country has significant inequalities, exhibiting a Gini index of 0.553 of 2023 (the Gini index determines a nation’s level of income inequality by measuring the income distribution or wealth distribution across its population) [27].

Analytical, ecological, observational study with repeated measures based on data collected from several national sources, including: the Bank of the Republic (central bank of Colombia), the National Ministry of health, the National Administrative Department of Statistics (DANE), the National Planning Department (DNP), and the National Health Institute (INS) [24,26,28]. The study population included the entire population with dengue registered cases at the municipal level during the specified period. Monthly data from January 2015 to December 2020 were analyzed, covering a total of 1,110 municipalities in Colombia.

### Data collection of Dengue data

Data on confirmed cases of dengue fever in the 32 departments and municipalities of Colombia were obtained from the National Public Health Surveillance System (SIVIGILA) [29], based on the ICD-10 registry definition of dengue case (A90: Classic dengue and/or A91 Dengue hemorrhagic fever) established by healthcare institution as either confirmed with laboratory testing or a probable case of dengue fever with clinical warning signs such as headache, retro ocular pain, myalgia, arthralgia, rash or rash and also has a history of displacement (up to 15 days before the onset of symptoms), severe and continuous abdominal pain, persistent vomiting, diarrhea, drowsiness and/or irritability, postural hypotension, painful hepatomegaly >2cms, decreased diuresis, drop in temperature, mucosal bleeding, abrupt drop in platelets (<100.000) associated with hemoconcentration or residing in a dengue endemic area [30]. The data used in this study was extracted from this system: the authors gathered all the weekly dengue reports at a municipal/local level based [24,25]. For the analysis, the data were aggregated into monthly case counts per municipality to facilitate temporal and spatial comparisons.

### Socioeconomic, geographic, climatic, and infrastructure data

Sociodemographic and economic variables included population density (inhabitants/km²) and educational coverage (%), obtained from the National Administrative Department of Statistics (DANE) [31]; the multidimensional poverty index (MPI) and government-subsidized health coverage (%), obtained from the National Planning Department (NPD) through its Terridata platform [32]; and GDP per capita (millions of dollars), obtained from the Central Bank [33]. On the other hand, infrastructure and access to public services indicators, such as aqueduct coverage (%), education coverage (%), healthcare coverage subsided by the government (%), and access to public services (%), were also obtained from the Terridata platform [32]. Regarding to governance variables, included the Municipal Performance Measure (%) defined as a composite indicator developed by the National Planning Department of Colombia that evaluates the efficiency and management capacity of local governments, covering administrative, fiscal, and planning dimensions (%) were obtained from Terridata platform of NDP [32] while Multidimensional poverty index (MPI) incidence (%) were obtained from the National Administrative Department of Statistics (DANE) [31].

Geographic and cartographic information were obtained from the DANE Geoportal public website [34]. Climatic and environmental data were provided by World Bank Group (WBG) in the Climate Change Knowledge Portal (CCKP), what is a hub for climate-related information for the WBG [35] or purposes of provides an online platform from which access and analyze comprehensive data related to climate change and development, [35] from this platform were obtained data from monthly maximum temperatures, monthly minimum temperatures, monthly relative humidity and total monthly rainfall. On the other hand, the National Oceanic and Atmospheric Administration (NOAA) provided information of the ENSO phenomenon by the period of analysis [36].

Most variables were collected on a monthly or annual basis depending on their reporting frequency and were available at the municipal level, except for GDP per capita and climatic variables, which were reported at the departmental level.

### Statistical analysis

Statistical analysis was conducted using R 4.2.0 and Stata 17MP software. About the descriptive analysis, absolute and relative frequencies were calculated for the qualitative variables. Measures of central tendency (average and median) were estimated for the quantitative variables. Standard deviation and interquartile range were assessed for the dispersion measures. These analyses were performed stratified by the years of analysis included in the study.

For purpose of the analysis, all municipalities with at least one case reported of dengue in the sample were included. With the aim of comparing number of cases of dengue reported during the study analysis period at the territorial level, we visualized the spatiotemporal patterns of dengue in Colombia, using choropleth maps based on quantiles groups, using ‘spmap’ command in Stata for this purpose. The geographic base layer used for the maps (Colombian municipal boundaries) was sourced from the publicly available shapefile provided by the DANE geoportal. The shapefile used is available from: https://geoportal.dane.gov.co/

To analyze the relationship among socioeconomic, geographic, climatic, and infrastructure data factors with number of cases reported in the country between 2015-2020 data was analyzed using a Generalized Additive Mixed Model (GAMM), the estimations were conducted using the ‘mgcv’ package. A GAMM model was estimated to study the expected number of cases 𝑌_𝑖𝑗𝑡_ for each territory 𝑖 in each time 𝑗, assuming that 𝑌_𝑖𝑗𝑡_ ∼ 𝑁𝑒𝑔𝐵𝑖𝑛(𝜇_𝑖𝑗𝑡_, 𝜃), with 𝜃 the dispersion parameter of the negative binomial distribution, based on 𝑘 variables as follows:

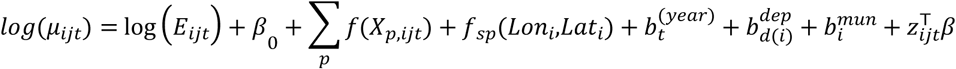

Where 𝐸_𝑖𝑗𝑡_ is the municipal population, 𝑧_𝑖𝑗𝑡_ is the effect of categorical variables (ENSO, municipality category), 𝑓_𝑝_(.) are smooth functions for the variables included in the analysis in the time 𝑗 trend defined via penalized cubic regressions splines; 𝑓_𝑠𝑝_(𝐿𝑜𝑛,𝐿𝑎𝑡) is the two-dimensional spatial term 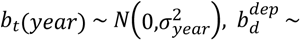 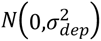 and 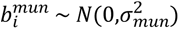 are time-specific and department-specific randoms effects to capture the effects of potential unobserved confounders in the model. Considering the high level of dispersion in the number of dengue cases, a negative binomial model was estimated based on the fast restricted maximum likelihood method (fREML). In order to adjust the models to the underlying population at risk, we included the annual municipal population as an offset term in the models, to estimate rates based on the number of events that occur in a defined population during a specified period, divided by the population at risk. In a negative binomial model linked to the logarithm, this compensation limits its coefficient to 1, ensuring that the model estimates rates rather than absolute counts.

The generalized cross-validation or GCV criterion was used to select an appropriate smoothing parameter value for the proposal models. Given the possible spatial autocorrelation structure, a spatial Generalized Additive Mixed Model (spatial GAMM) was fitted to evaluate the association between dengue cases and sociodemographic, climatic and performance government factors accounting for spatial and temporal structure. The model included a smooth function of the MVI, a two-dimensional spline over geographic coordinates (longitude and latitude), and a random effect for year.

The effect of sociodemographic, climatic and performance government variables were adjusted using the following confounding variables identified in the directed acyclic diagram (DAG, see supplementary material Fig. S1): altitude, population density, aqueduct coverage, education coverage, subsidized survey coverage, public services coverage, municipal performance measurement, precipitation, maximum and minimum temperature, relative humidity, Multidimensional Poverty Index (MPI) and GDP per capita. These variables were used as covariates, based on previous literature, what reported and the effect of the climate variables on dengue fever, the impact of socioeconomic conditions on dissemination of vector-borne diseases and evidence associated with the role of performance institutional in the control of *Aedes*.

Several models were built to adjust the sociodemographic, climatic and performance government variables according to the multilevel component of the information, the performance of these models was evaluated using Akaike’s Information Criterion (AIC) and Bayesian Information Criterion (BIC) and ANOVA test. The smallest AIC/BIC indicates the model with the best fit. Moreover, a diagnostic analysis of the models was performed based on simulated residual diagnostics from the ‘DHARMa’ R package [37], evaluating residual uniformity (Kolmogorov-Smirnov test), overdispersion, and outliers. Additionally, QQ plots and residuals versus predicted plots were inspected to evaluate potential systematic deviations. The spatial GAMM estimations were conducted using the ‘mgcv’ R package [38], applying penalized cubic regression splines to allow for flexible smoothing of the relationship between selected variables and dengue occurrence over time. Statistical analysis was conducted using R 4.1.1 and Stata 17 MP software.

## Results

### Study population

A total of 1,100 municipalities were included in the study. Table 1 describes the characteristics of these territories. The median altitude above sea level was 1,010 meters (range: 1.00-3,850 meters). The median values of health and education coverage indicators were above 50%, contrasting with the median water supply coverage, which was below this value. The trend of dengue cases is illustrated in Figure 1. During the study period, a total of 421798 dengue cases were reported in Colombia. At the national level, the annual number of cases showed marked interannual variation, with a median of 69688 cases per year (IQR: 43820–95288) and a peak of 118956 cases in 2019. Across municipalities, the annual median number of reported cases was 6280 (IQR: 2842–9278), reflecting the high spatial heterogeneity of dengue transmission. The highest numbers of cases were reported in 2019 (118956) and 2016 (91378) (red color in Fig 1), whereas the lowest occurred in 2017 (24448). (blue color in Fig 1). Between 2015 and 2020, dengue incidence showed marked interannual fluctuations, peaking in 2015 and 2019. Socioeconomic and municipal performance indicators improved steadily, while the multidimensional poverty index decreased over time. These results are summarized in Table 1.

**Fig 1.**
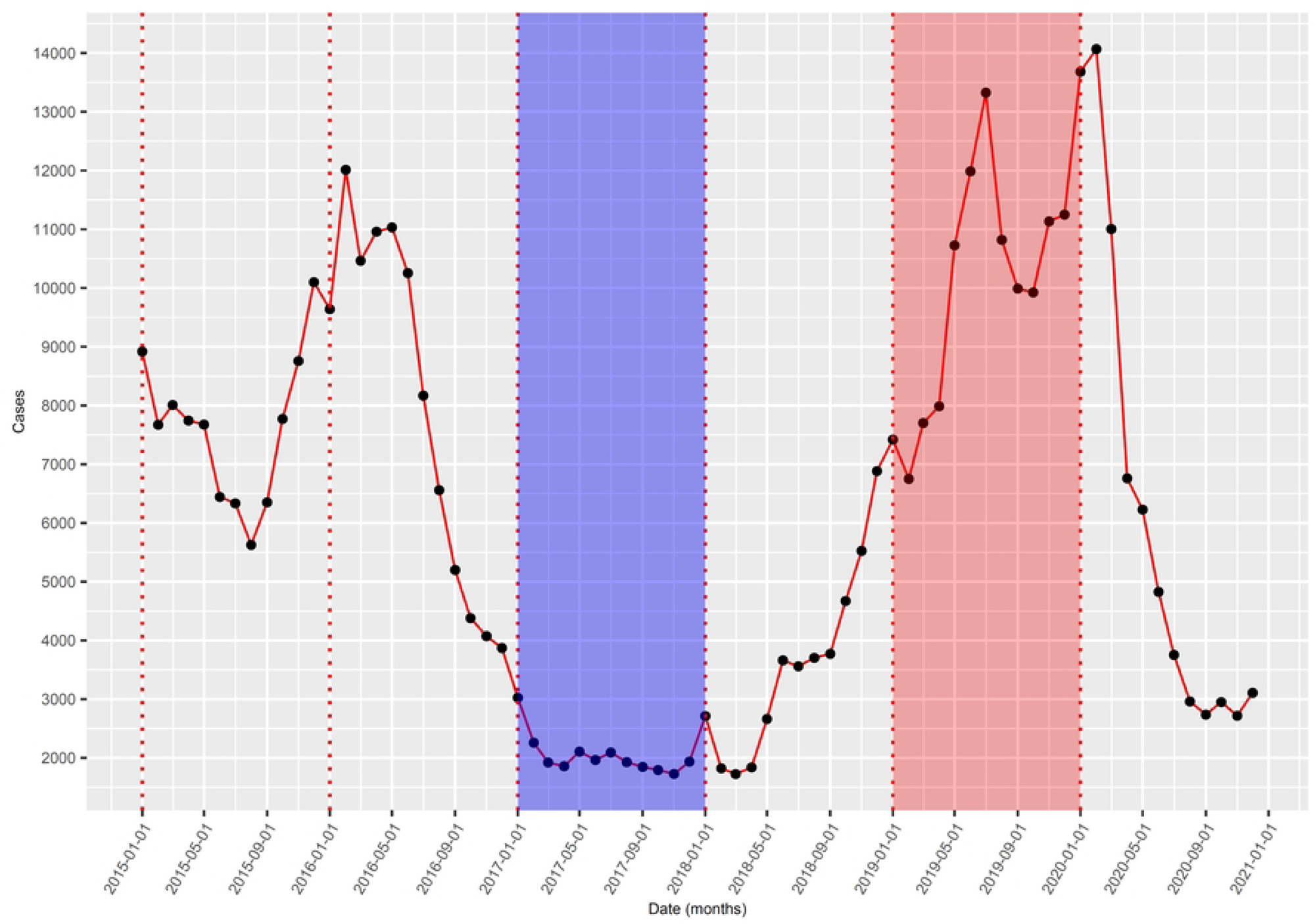
Total number of cases per year of dengue in Colombia.

**Table 1.** Table of sociodemographic characteristics of the study population.

| Variables <sup>a</sup> | 2015 | 2016 | 2017 | 2018 | 2019 | 2020 |
| --- | --- | --- | --- | --- | --- | --- |
| Total of reported cases of dengue fever | 91378 | 96592 | 24448 | 42427 | 118956 | 47997 |
| Reported cases of dengue fever (median [IQR]) | 7.00 [0.00-42.00] | 7.00 [0.00-36.00] | 2.00 [0.00-9.00] | 1.00 [0.00-14.00] | 7.00 [0.00-66.50] | 6.00 [0.00-35.00] |
| Annual incidence dengue rate (per 100000 population) (median [IQR]) <sup>b</sup> | 53.94 [0.00-238.10] | 50.50 [0.00-205.46] | 11.38 [0.00-53.29] | 12.20 [0.00-75.61] | 56.73 [0.00-322.81] | 35.08 [0.00-164.40] |
| Height above mean sea level (mean (SD)) | 1087.06 (901.05) | 1089.78 (902.48) | 1089.78 (902.48) | 1090.73 (902.34) | 1089.78 (902.48) | 1088.87 (917.08) |
| Population density (hab/km<sup>2</sup>) (median [IQR]) | 43.52 [20.31-87.80] | 43.76 [20.28-88.31] | 43.99 [20.32-88.17] | 44.36 [20.49-89.66] | 45.48 [20.79-91.63] | 43.63 [20.68-89.95] |
| Coverage in aqueduct (%) (mean (SD)) | 60.21 (29.51) | 57.70 (28.90) | 59.07 (28.39) | 69.86 (22.36) | 60.24 (28.22) | 60.34 (28.26) |
| Sewerage coverage (%) (median [IQR]) | 41.48 (29.07) | 40.04 (28.14) | 40.50 (27.26) | 46.27 (24.96) | 41.97 (27.56) | 42.31 (27.29) |
| Education coverage (%) (median [IQR]) | 83.69 [72.21-95.00] | 83.23 [71.83-95.68] | 82.54 [70.68-95.67] | 81.66 [69.67-96.25] | 88.42 [80.26-96.07] | 87.64 [79.68-94.96] |
| Subsidized regime coverage (%) (median [IQR]) | 97.60 [96.69-98.30] | 98.64 [97.84-99.12] | 98.94 [98.15-99.33] | 98.42 [97.68-98.90] | 99.36 [98.74-99.62] | 98.86 [98.13-99.26] |
| Transparency of the royalty project management index (%) (median [IQR]) | 73.66 [62.22-86.44] | 73.64 [62.19-86.44] | 83.80 [64.09-95.50] | 86.43 [67.80-96.53] | 85.87 [64.64-97.07] | 85.19 [65.74-96.48] |
| Access to public services (%) (mean (SD)) | 45.78 (11.81) | 45.77 (11.80) | 46.82 (11.36) | 46.86 (11.78) | 47.24 (11.06) | 48.01 (11.05) |
| Municipal Performance Measurement (%) (mean (SD)) | 47.84 (9.68) | 47.83 (9.67) | 49.03 (9.69) | 50.18 (9.33) | 54.40 (10.62) | 50.36 (11.11) |
| Multidimensional poverty index (%) (mean (SD)) | 41.35 (10.96) | 41.34 (10.95) | 36.76 (13.61) | 39.90 (19.97) | 47.05 (16.15) | 35.47 (15.22) |
| GDP per capita (USD millions) (mean (SD)) <sup>c</sup> | 5709.96 (2268.61) | 5407.33 (1902.93) | 5844.88 (2128.25) | 6151.59 (2360.89) | 5804.96 (2261.13) | 4663.05 (1691.96) |
| Precipitation (mm/month) (median [IQR]) | 154.95 [128.11-201.78] | 181.18 [157.17-242.37] | 165.86 [133.89-207.05] | 153.39 [126.75-215.59] | 149.83 [122.38-220.80] | 153.39 [126.83-215.56] |
| Maximum temperature (°C) (mean (SD)) | 27.45 (3.42) | 27.31 (3.48) | 26.86 (3.51) | 26.78 (3.45) | 27.23 (3.43) | 27.25 (3.50) |
| Minimum temperature (°C) (mean (SD)) | 18.59 (4.03) | 18.75 (3.88) | 18.47 (3.89) | 18.36 (3.89) | 18.70 (3.90) | 18.56 (3.95) |
| Relative humidity (%) (mean (SD)) | 79.36 (5.25) | 80.68 (4.63) | 80.81 (4.51) | 80.67 (4.64) | 80.17 (5.04) | 79.48 (4.94) |

a. Data are presented as mean (SD) for approximately symmetric variables and median [IQR] for skewed variables.
b. Annual dengue incidence rate was calculated as the total number of reported dengue cases per municipality per year divided by the mid-year population, multiplied by 100000.
c. Based on mean TRM for each year

Figure 2 shows the spatial and temporal patterns in the incidence of dengue for all municipalities in Colombia during 2015–2020. Most cases occurred in the north and southwest coast region, with the highest incidence in the Caribbean region. The frequency in southwest part of the coast region and eastern region increased in 2016 and 2020 compared with that in the previous years.

**Fig 2.**
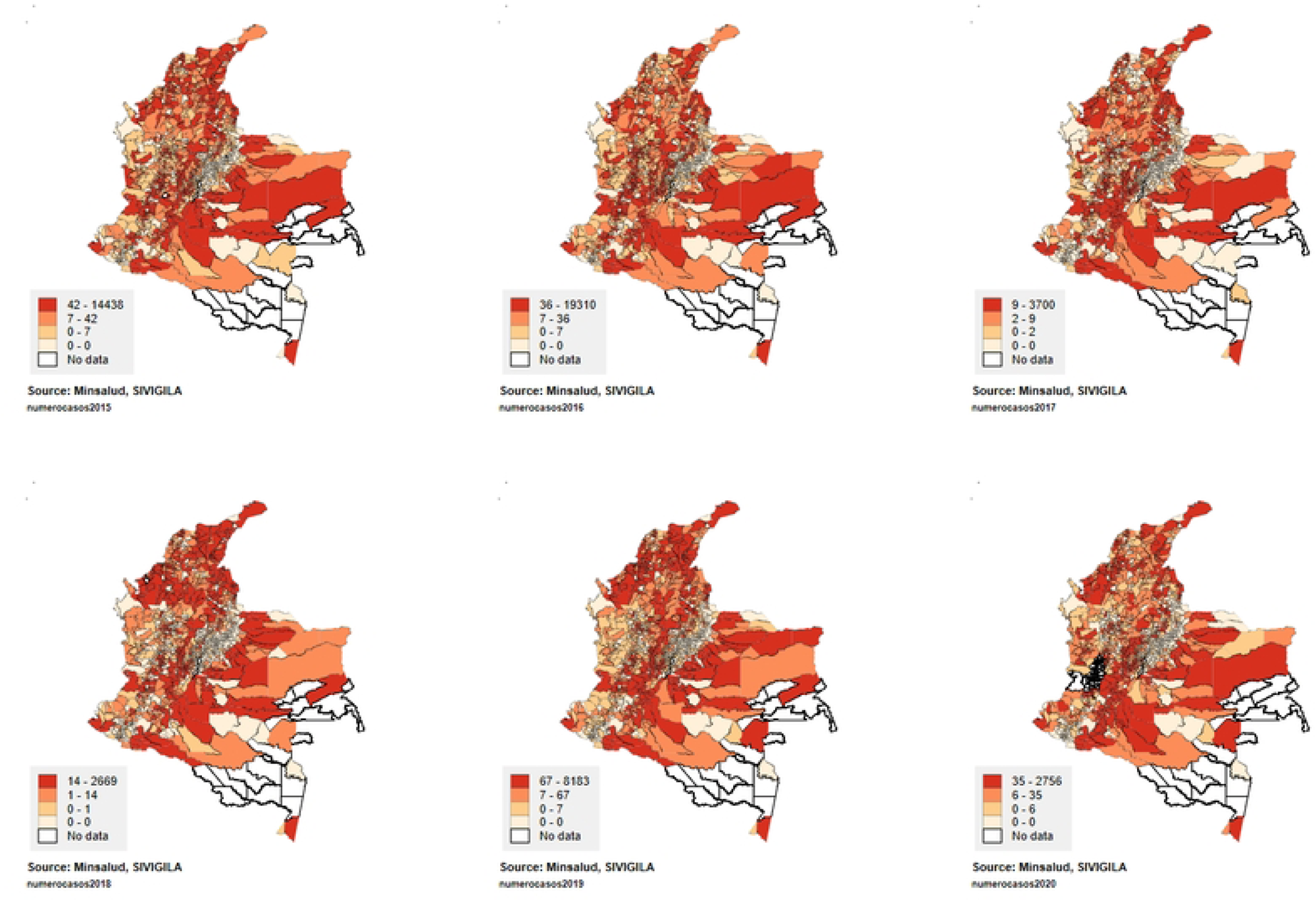
Spatial distribution of reported cases of dengue fever in the study period

### Spatial GAMM Analysis

Table 2 presents the estimates of the Negative Binomial Spatial GAMM for the number of dengue cases reported. Final model was selected after comparing alternative specifications, including models with fixed effects for department and year, models without the spatial component, and models with different random effect structures. Model selection was based on Akaike’s Information Criterion (AIC) and Bayesian Information Criterion (BIC), with the chosen specification yielding the lowest AIC and BIC values. The selected model explained 54% of the deviance in dengue cases. The high values of the effective degrees of freedom (edf) for the smooth functions indicate strong nonlinear associations between dengue incidence and sociodemographic, climatic, and governance-related variables; most of these findings were statistically significant.

**Table 2.**
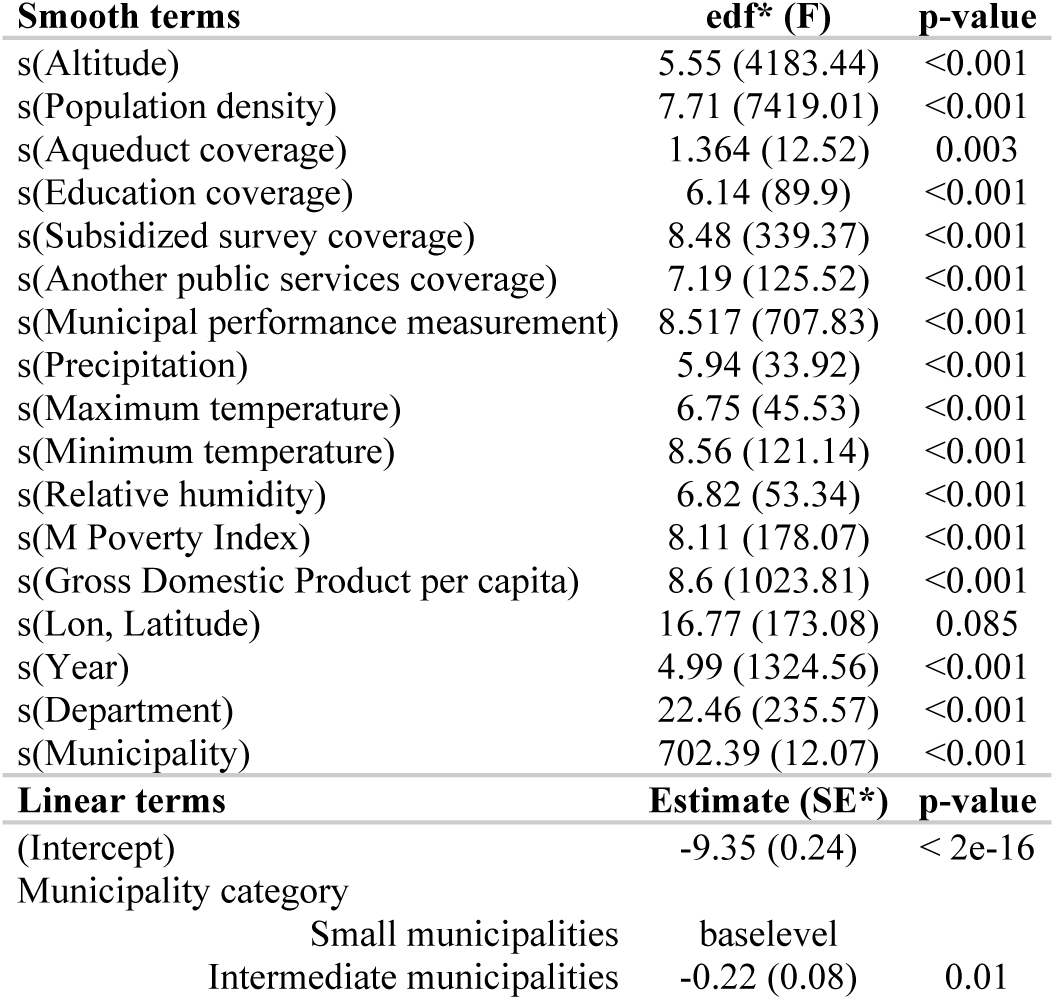
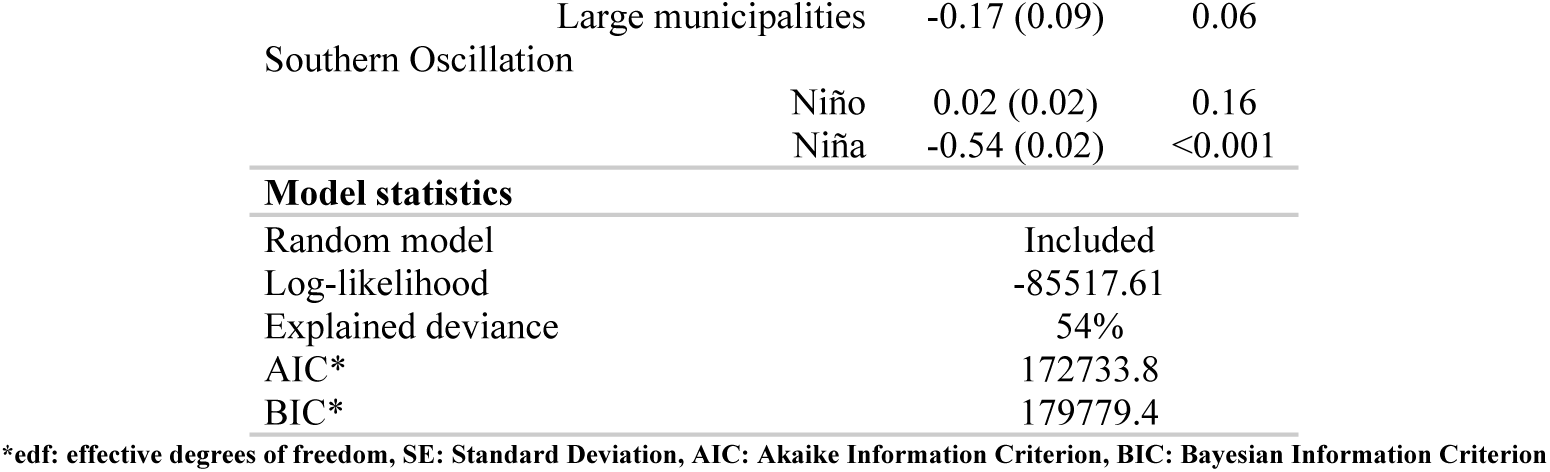
Model Spatial GAMM estimates of the effect of climatic, sociodemographic and performance conditions on Dengue disease in Colombia.

Figures 3, 4 and 5 show the relationships estimated by the final model. Altitude showed a multimodal pattern, with higher risk at low and medium altitudes (<750 m.a.s.l), a reduction around 1000–2800 m.a.s.l, and an uptick toward >3000 m.a.s.l (Figure 3a). Population density displayed a bimodal relationship, with peaks around 2000 and 10,000 P/km², followed by declines at higher levels (Fig. 3b). Multidimensional Poverty Index (MPI) showed irregular oscillations without a consistent gradient, although incidence appeared higher in territories with 35–65% of people in multidimensional poverty (Fig. 3c). Finally, GPD per capita (Fig. 3d) show directly proportional trends, where dengue fever is concentrated in areas with high GDP levels, particularly concentrated in territories above 9000 USD.

**Fig 3.**
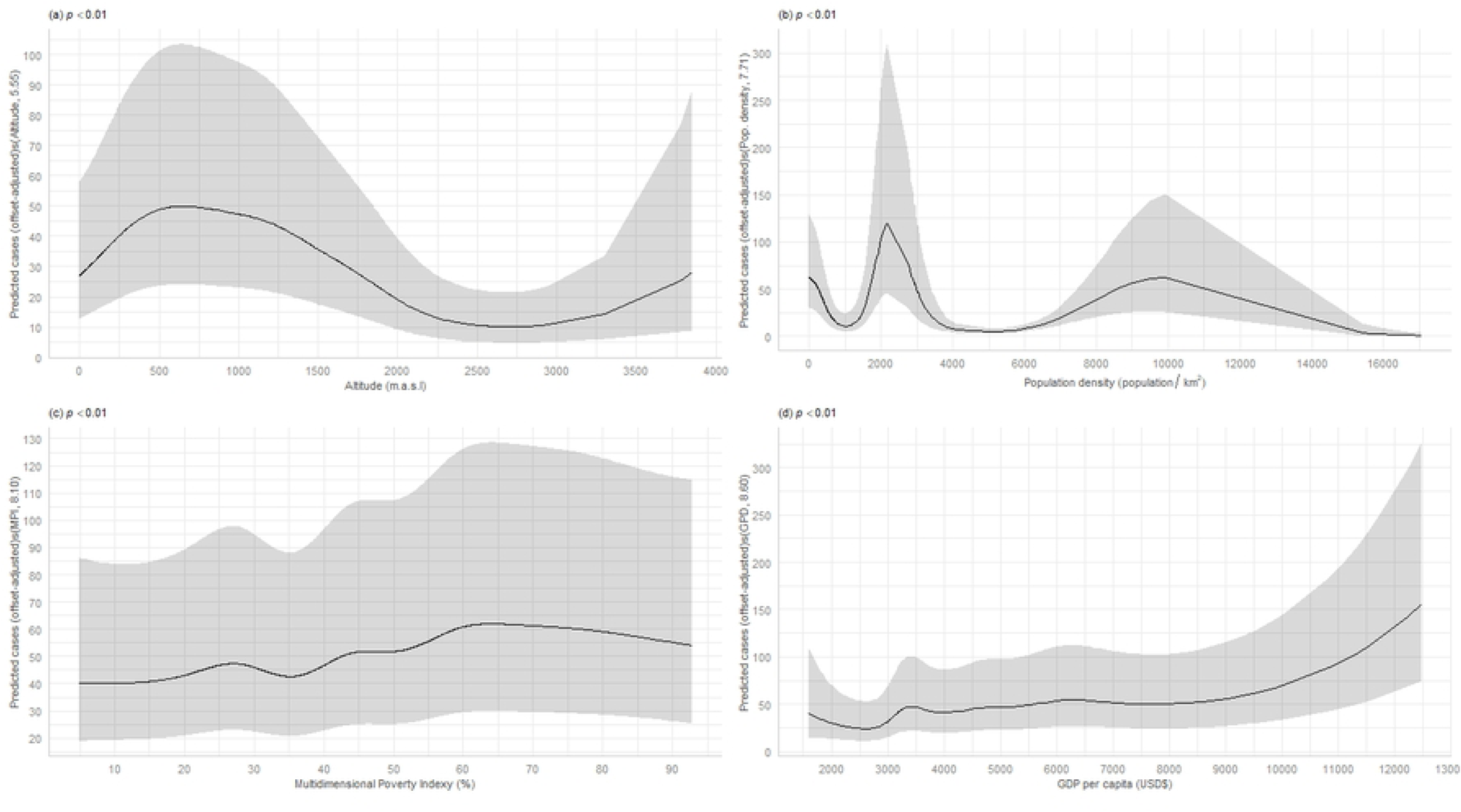
Spatial Generalized Additive Mixed Models-estimated relationships between nonlinear geographic and socioeconomic factors and dengue cases in Colombia based on data from all years 2015−2020.

**Fig 4.**
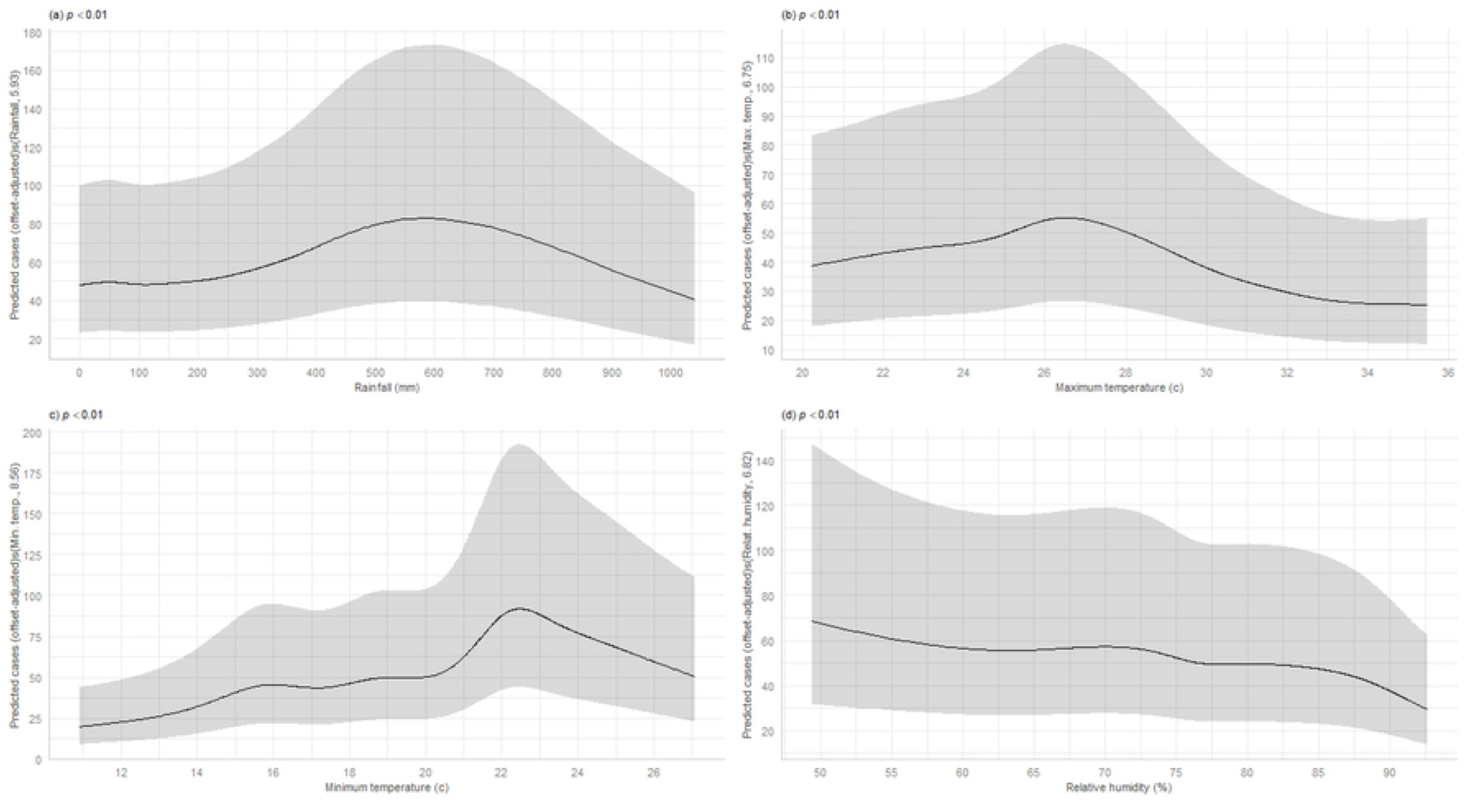
Spatial Generalized Additive Mixed Models-estimated relationships between probable nonlinear climatic factors and dengue cases in Colombia based on data from all years 2015−2020.

**Fig 5.**
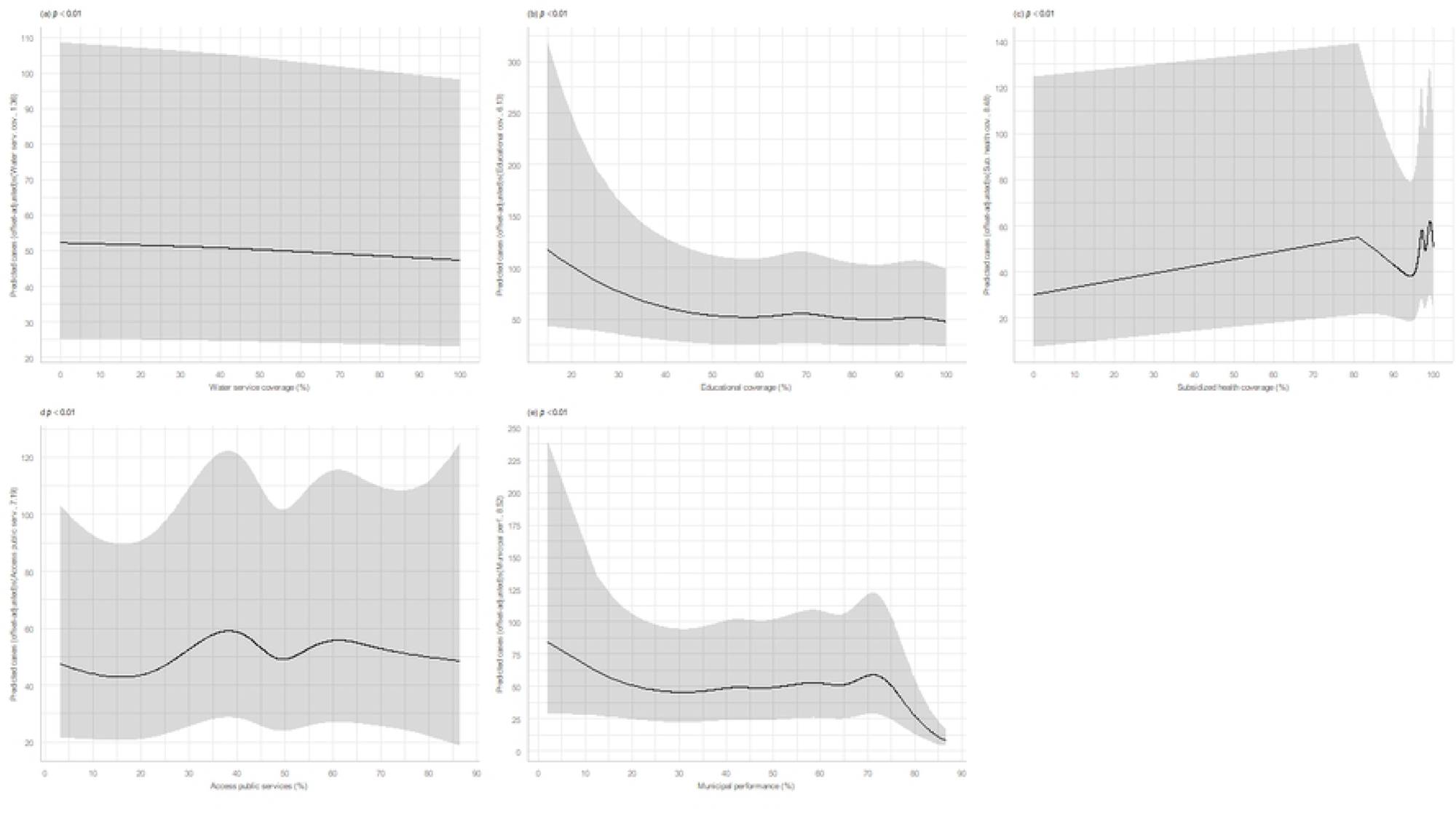
Spatial Generalized Additive Mixed Models-estimated relationships between potentially nonlinear infrastructure factors and dengue cases in Colombia based on data from all years 2015−2020.

Figure 4 shows the climatic component in the analysis: rainfall was associated with the highest incidence around 500–700 mm. Maximum and minimum temperatures presented unimodal associations, with higher incidence at intermediate ranges (25–28 °C and 21–24 °C, respectively), and lower incidence at extreme values (Figs. 4a, 4b, 4c) showing an increase in dengue frequency towards the middle of the distribution and gradually decreasing at that point. Relative humidity showed a negative, nearly linear association, with decreasing dengue occurrence at higher humidity levels (Fig. 4d).

Figure 5 shows the government performance in the analysis, water service coverage exhibited a negative, linear pattern, with lower incidence at higher coverage (Fig. 5a). Educational coverage showed a negative relationship (Fig. 5b), whereas subsidized health coverage showed a positive association, albeit with wide uncertainty at extreme values (Fig. 5c). Access to public services displayed a bimodal pattern, with higher incidence at intermediate levels (40–70%) and lower incidence at higher levels (Fig. 5e). Finally, municipal performance showed a marked negative relationship, with better-performing municipalities consistently exhibiting lower reported dengue cases (Fig. 5f).

On the other hand, differences between municipality category (Small versus Intermediate municipality: p value<0.01) and ENSO dynamic (p value<0.01) were found in the frequency of dengue for the period of analysis (Table 2). These patterns suggest that the level of environmental, socioeconomic, and governmental vulnerability jointly influence the risk of dengue in Colombia, highlighting that climatic factors alone do not fully explain incidence trends.

The DHARMa diagnostics indicated adequate dispersion (p = 0.108), but significant deviations from uniformity and presence of outliers (p<0.05). Nonetheless, residuals vs. predicted plots showed no major systematic biases, supporting overall model adequacy (Fig. 6).

**Fig 6.**
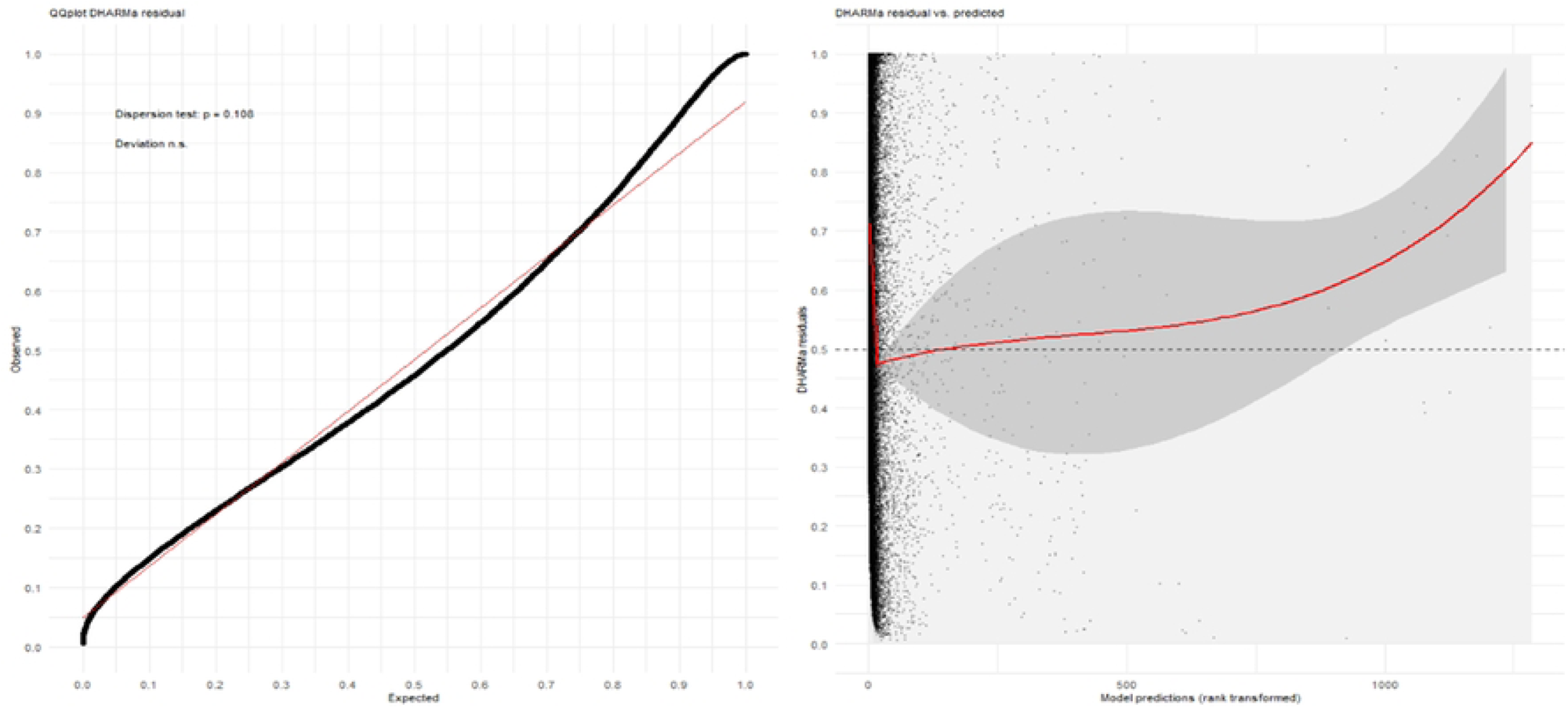
Residual Diagnosis. Left is the Q-Q plot for the DHARMa residuals whereas the relationship between the reported and predicted cases is shown on Right.

## Discussion

This study aimed to determine the relationship between the frequency of dengue fever and socioeconomic, geographic, climatic, and infrastructure data factors in Colombia between January 2015 and December 2020. The interannual variation in dengue incidence, with peaks in 2015 and 2019, aligns with previously reported national epidemic cycles in Colombia. Given the complex interaction between the socioeconomic, geographic, and infrastructure factors and the dynamics of dengue, spatial GAMM models provide more flexible modeling to analyze the spatiotemporal dynamics of dengue, allowing the inclusion of non-linear relationships of the different factors included in the model. [39]. The results showed that the incidence of dengue is strongly associated with climatic factors, with a nonlinear association between rainfall and temperature, and with complex socioeconomic and infrastructure conditions. The spatial GAMM approach allowed us to capture temporal and spatial structure relationships and reduce residual spatial autocorrelation compared to conventional models.

Access to public services, such as water, was associated with a decrease in dengue case frequency, which may highlight the need for improved access to basic services in affected communities as suggested by prior studies [40,41]. Moreover, the study highlighted the links between multidimensional poverty index and dengue incidence, with higher prevalence in areas with higher poverty levels in line with prior studies describing this association [42,43]. The study also identified education and information access as crucial factors in dengue prevention and control, along with an increase in the GDP. Similarly, a prior study described that education and access to information are crucial for dengue prevention and control, and gains in GDP can reinforce these measures by improving living conditions, health infrastructure, and vector control capacity [44]. On the other hand, the results show a significant influence of climatic components (temperature, humidity, and rainfall) on the frequency of dengue in Colombia. Precipitation showed a unimodal association, with a higher incidence around 500-700 mm, which may be related with the optimal conditions for *Aedes* reproduction due to water storage in containers and the accumulation of standing water, while very heavy rainfall can wash away breeding sites, reducing mosquito populations [45,46]. Maximum and minimum temperatures also showed concave associations, with the highest risk in the range of 26-28 °C and 22-24 °C, respectively, consistent with the optimal temperature ranges for mosquito survival and viral replication described in the literature [47,48]. These non-linear relationships may be explained by the biology of the vector, as higher precipitation rates combined with higher temperatures result in increased humidity, which is associated with an increase in the feeding activity, survival, and egg development of *Aedes aegypti* [49].

Likewise, temperatures play a crucial role in vector dissemination, as *Aedes* mosquitoes depend on specific temperature ranges between 21°C and 31°C for survival and reproduction [50,51]. Precipitation also plays a fundamental role in increasing dengue incidence. Rainfall ranges between 400mm and 500mm increase the risk of dissemination by creating new reservoirs that allow vector oviposition in natural environments [46]. The relationship with relative humidity was negative, showing that high values reduce the incidence of dengue, possibly due to lower survival rates of eggs and larvae in saturated environments [46]. These conditions of humidity, precipitation, and temperature make the presence of *Aedes* widespread in tropical and subtropical environments. In these regions, temperature and humidity levels, combined with prolonged rain cycles, create an ideal environment for the mosquito’s life cycle [52]. These findings are consistent with other Latin American studies, highlighting the importance of climatic control and surveillance in territories most vulnerable to dengue dissemination in the context of climate change [53,54]. The impact of climatic vulnerability on dengue risk is also consistent with results from other global studies [55].

Regarding sociodemographic and governance-related vulnerabilities, strong non-linear relationships were observed with government performance aspects, water service coverage, sewerage, education, and health coverage. For water supply, a polynomial trend was observed rather than a decrease in dengue frequency in territories with higher coverage. This finding contrasts with previous evidence that showed an inverse relationship, with higher dengue dissemination in areas with less water access due to increased artificial reservoirs from water storage [56]. This implies an increase in breeding sites for mosquitoes, both in natural puddles formed by rain and in household water storage in areas affected by heat waves [18,57]. However, this effect can be modified by other factors such as inequity, income, and ineffective public health policies [50]. The effects of socioeconomic determinants in various Latin American countries highlight the biophysical, political-institutional, and community risks for dengue dissemination, addressing marginalization and inequity issues in primary health care and their consequences in terms of living conditions, access to public services, and education [58]. These findings are similar in other regions of the world, where the study of social inequality, poverty, and inequity effects on dengue dissemination suggests integrated efforts beyond conventional vector control strategies, aiming to control the dengue threat through improvements in socioeconomic conditions, income, and education. [59,60].

The approach of assessing climatic vulnerabilities jointly with social and economic factors is crucial to inform academia and policymakers with valuable information that facilitates climate adaptation and coping strategies at various scales [17,61,62]. The degree of vulnerability to climate change is differential and attributable to households’ socioeconomic variations and access to basic services [61,63]. Considering these data and the global spread of the *Aedes Aegypti* [57,64], vector control emerges as one of the most effective methods to mitigate new dengue epidemics. However, this measure has proven insufficient [65,66]. Evidence suggests implementing integrated preventive strategies to eliminate breeding sites, educate at-risk populations, disseminate climate information, and identify territories with high socioeconomic and climatic vulnerability. [67]. In the context of accelerating climate change, the nonlinear climate-dengue relationships identified in this analysis may have important implications. The unimodal associations suggest that both increases and decreases in precipitation or temperatures beyond optimal ranges could paradoxically reduce transmission in some areas while increasing it in others. However, changes in climate variability and extreme events may have effects beyond those captured by monthly averages. Future research should explore how climate change projections interact with socioeconomic development pathways to forecast dengue risk.

By incorporating spatial smoothing and random effects for year and department, spatial GAMM provided several advantages over conventional regression, as mentioned in the literature in terms of modeling complex relationships, predictive power, geographic connectivity, and model variations in true disease pattern [73]. It allowed flexible modeling of non-linear associations without imposing restrictive assumptions, accounted for residual spatial autocorrelation, and captured heterogeneity across territorial analysis units [68]. This approach complements earlier ecological studies that relied on linear or Poisson models, providing a more nuanced understanding of the climatic and social risk for dengue transmission. These methodological advances can support more accurate local predictions and better-targeted interventions. Moreover, the results presented in this study may have important implications for dengue management in Colombia and similar endemic contexts. Monitoring and analyzing dengue transmission windows presents opportunities for a climate-based early warning system, especially during “El Niño” seasons, when frequency was higher. Secondly, the nonlinear relationships with infrastructure and governance suggest that investment in improving water or sanitation services may not be sufficient and that community-level interventions are needed to mitigate risk. On the other hand, findings related to education coverage and local administration highlight the need to strengthen institutional capacity and community participation as part of integrated vector management strategies. Finally, the relationship with GDP highlights the importance of focusing not only on the most vulnerable territories, but also on major urban areas with high population density and excessive growth, where the risk of dengue remains high despite greater economic development.

Our study had limitations, first, a significant limitation is related to underreporting bias in health statal databases regarding inequities in access to basic services, which has been described in the literature and can be as high as 5% [69]. Additionally, the municipal-level disaggregation of healthcare system affiliation variables can be considered a limitation in this analysis due to difficulties in obtaining more precise data on the existing gaps between the contributory and subsidized regimes. Furthermore, we could not account for population mobility between municipalities, which affects both exposure patterns and case reporting locations. Lack of dengue serotype data limited assessment of viral strain dynamics, and absence of entomological surveillance data prevented direct vector-climate validation. On the other hand, given the ecological design of this study, inference at the individual level cannot be possible and raises the possibility of ecological fallacy. Associations observed at the municipal level may not reflect individual-level relationships, and ecological fallacy remains a possibility. The cross-sectional nature of the aggregated data prevents establishment of temporal precedence for many relationships. Dengue surveillance data may also be affected by underreporting and variability in diagnostic capacity across municipalities. The use of monthly case aggregation may mask within-month transmission dynamics, and the optimal lag structures between climatic exposures and disease outcomes may vary across municipalities. Residual confounding by housing quality and sanitation practices cannot be excluded. Moreover, we used annual population projections as the denominator for incidence estimation, while cases were aggregated monthly. More studies to better understand these dynamics and develop effective interventions to reduce dengue transmission in Colombia. Despite these limitations, the large dataset, spatially explicit modeling, and sensitivity analyses provide confidence in the robustness of the findings.

Overall, the combination of climatic factors, service coverage, municipal performance, and transparency of social development presents an integral scope that reflects the complexity of the challenges Colombia faces in the fight against dengue. Our findings suggest that disparities in access to essential services and climatic variability are intrinsically linked to the prevalence of dengue.

## Conclusion

This multidimensional study on dengue in Colombia between 2015 and 2020 reveals a complex interaction between socioeconomic, geographic, climatic, and infrastructure factors. Coverage of public services such as water was found to be associated with a decrease in dengue occurrence, while multidimensional poverty showed more intricate relationships, with higher incidence in areas with higher levels of poverty. Furthermore, increased education and access to information were highlighted as crucial factors, along with an increase in GDP. These findings highlight the need for comprehensive approaches in dengue prevention and control policies, adapted to the specific conditions of each community. More research is required to better understand the dynamics of dengue and develop effective interventions that reduce its burden on the population. Furthermore, factors identified at the local level can be useful predictors at the regional level, which can help anticipate and address vulnerable populations on a broader scale, especially in the context of climate change.

## Data Availability

The dengue surveillance data analyzed in this study were obtained from Colombia’s National Public Health Surveillance System (SIVIGILA) and are available through the National Health Institute (INS) upon reasonable request subject to data sharing agreements. Socioeconomic and municipal performance data are publicly available from the National Planning Department (DNP) Terridata platform (https://terridata.dnp.gov.co/). Climatic data are publicly available from the World Bank Climate Change Knowledge Portal (https://climateknowledgeportal.worldbank.org/). The geographic shapefiles are available from the Geoportal of the National Administrative Department of Statistics (DANE), Colombia’s National Statistics Office (https://geoportal.dane.gov.co/). License/terms of use: https://geoportal-dane-gov-co.translate.goog/acerca-del-geoportal/licencia-y-condiciones-de-uso/?_x_tr_sl=es&_x_tr_tl=en&_x_tr_hl=es&_x_tr_pto=wapp#gsc.tab=0. All analysis code is available from the corresponding author upon reasonable request.

## Data Availability

https://geoportal-dane-gov-co.translate.goog/acerca-del-geoportal/licencia-y-condiciones-de-uso/?_x_tr_sl=es&_x_tr_tl=en&_x_tr_hl=es&_x_tr_pto=wapp#gsc.tab=0

---

**S1 Fig.**
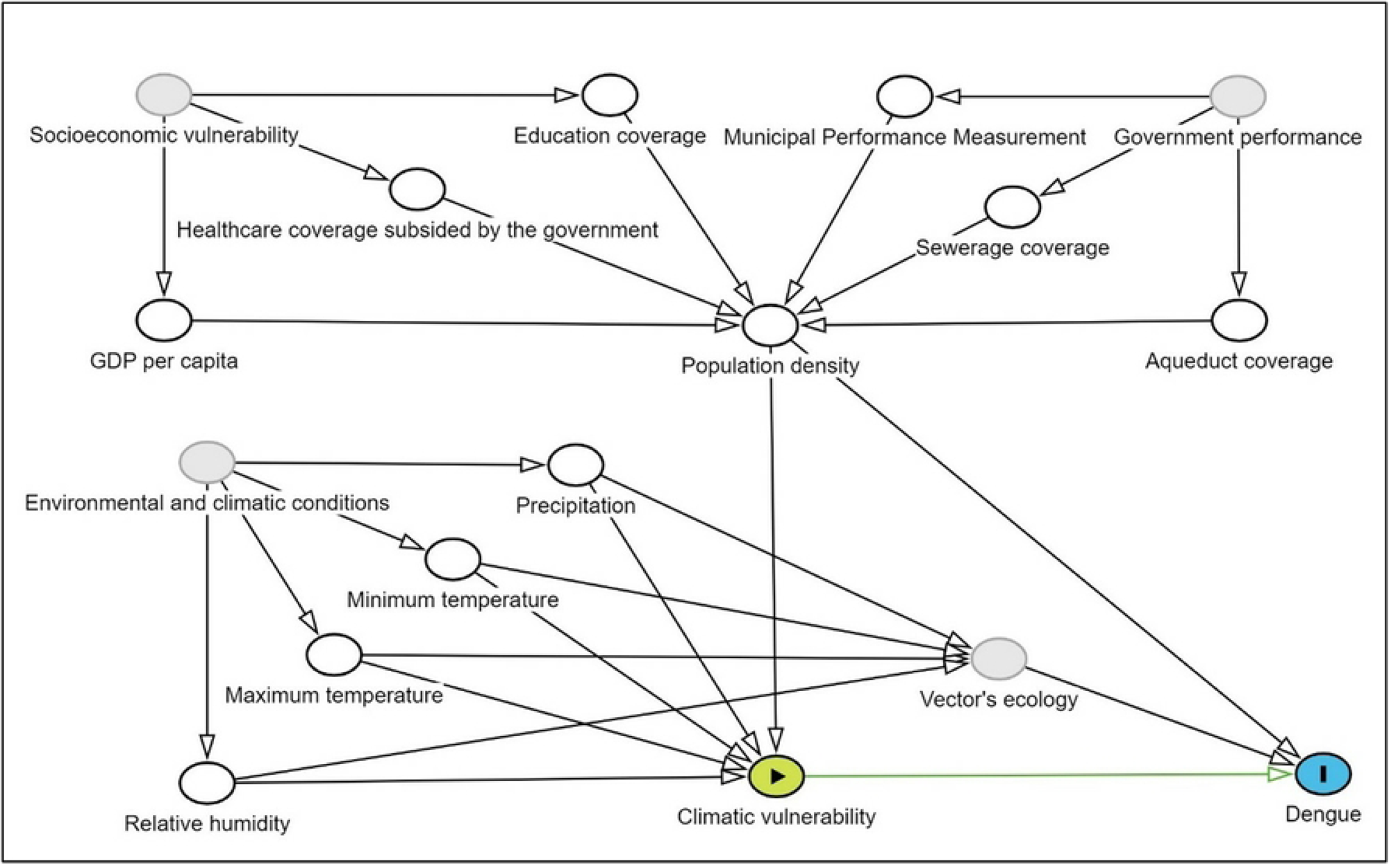
Directed acyclic diagram of the relationship dengue frequency and climatic, and sociodemographic and performance conditions.

## References

1. World Health Organization. Dengue control. In: WHO [Internet]. World Health Organization; 2017 [cited 22 Apr 2019]. Available: https://www.who.int/denguecontrol/epidemiology/en/

2. Organización Panamericana de la Salud. Casos de dengue superan los 1,6 millones en América, lo que pone de relieve la necesidad del control de mosquitos durante la pandemia. In: news [Internet]. 2020 [cited 28 Sep 2020] p. 1. Available: https://www.paho.org/es/noticias/23-6-2020-casos-dengue-superan-16-millones-america-lo-que-pone-relieve-necesidad-control

3. Instituto Nacional de Salud. Protocolo de vigilancia en salud pública Dengue. Bogotá; 2024. doi:10.33610/JQVP8800

4. Sutherst RW. Global change and human vulnerability to vector-borne diseases. Clin Microbiol Rev. 2004;17: 136–173.

5. Guo C, Zhou Z, Wen Z, Liu Y, Zeng C, Xiao D, et al. Global Epidemiology of Dengue Outbreaks in 1990–2015: A Systematic Review and Meta-Analysis. Front Cell Infect Microbiol. 2017;7: 317. doi:10.3389/fcimb.2017.00317

6. da Silva Oliveira LN, Itria A, Lima EC. Cost of illness and program of dengue: A systematic review. Borrow R, editor. PLoS One. 2019;14: e0211401. doi:10.1371/journal.pone.0211401

7. Brady OJ, Gething PW, Bhatt S, Messina JP, Brownstein JS, Hoen AG, et al. Refining the Global Spatial Limits of Dengue Virus Transmission by Evidence-Based Consensus. Reithinger R, editor. PLoS Negl Trop Dis. 2012;6: e1760. doi:10.1371/journal.pntd.0001760

8. Center for Disease Control and Prevention. Dengue. 2019 [cited 22 Apr 2019]. Available: https://www.cdc.gov/dengue/epidemiology/index.html

9. Global Burden of Disease data visualiation I for HM and. GBD Compare | IHME Viz Hub. In: Global Burden of Disease [Internet]. 2015 [cited 8 Oct 2019]. doi:http://ihmeuw.org/3pgz

10. Keating J. An investigation into the cyclical incidence of dengue fever. Soc Sci Med. 2001;53: 1587–97.

11. Kraemer MUG, Sinka ME, Duda KA, Mylne AQN, Shearer FM, Barker CM, et al. The global distribution of the arbovirus vectors Aedes aegypti and Ae. albopictus. Elife. 2015;4: 1–18. doi:10.7554/eLife.08347

12. Messina JP, Brady OJ, Pigott DM, Golding N, Kraemer MUG, Scott TW, et al. The many projected futures of dengue. Nat Rev Microbiol. 2015;13: 230–239. doi:10.1038/nrmicro3430

13. Bonifay T, Douine M, Bonnefoy C, Hurpeau B, Nacher M, Djossou F, et al. Poverty and Arbovirus Outbreaks: When Chikungunya Virus Hits More Precarious Populations Than Dengue Virus in French Guiana. Open Forum Infect Dis. 2017;4. doi:10.1093/ofid/ofx247

14. Pacheco-coral AP. The role of migration processes in dengue fever occurrence in Colombia : a mixed study approach. 2016.

15. Salas RN, Jha AK. Climate change threatens the achievement of effective universal healthcare. BMJ. 2019;366: l5302. doi:10.1136/bmj.l5302

16. Kloog I, Novack L, Erez O, Just AC, Raz R. Associations between ambient air temperature, low birth weight and small for gestational age in term neonates in southern Israel. Environ Health. 2018;17: 76. doi:10.1186/s12940-018-0420-z

17. Moreno-Banda GL, Riojas-Rodríguez H, Hurtado-Díaz M, Danis-Lozano R, Rothenberg SJ. Effects of climatic and social factors on dengue incidence in Mexican municipalities in the state of Veracruz. Salud Publica Mex. 2017;59: 41–52. doi:10.21149/8414

18. Barclay E. Is climate change affecting dengue in the Americas? Lancet. 2008;371: 973–974. doi:10.1016/S0140-6736(08)60435-3

19. Liu K, Sun J, Liu X, Li R, Wang Y, Lu L, et al. Spatiotemporal patterns and determinants of dengue at county level in China from 2005–2017. International Journal of Infectious Diseases. 2018;77: 96–104. doi:10.1016/j.ijid.2018.09.003

20. Rohat G, Monaghan A, Hayden MH, Ryan SJ, Charrière E, Wilhelmi O. Intersecting vulnerabilities: climatic and demographic contributions to future population exposure to Aedes -borne viruses in the United States. Environmental Research Letters. 2020;15: 084046. doi:10.1088/1748-9326/ab9141

21. Da Conceição Araújo D, Dos Santos AD, Lima SVMA, Vaez AC, Cunha JO, Conceição Gomes Machado de Araújo K. Determining the association between dengue and social inequality factors in north-eastern Brazil: A spatial modelling. Geospat Health. 2020;15. doi:10.4081/gh.2020.854

22. Udayanga L, Gunathilaka N. Climate change induced vulnerability and adaption for dengue incidence in Colombo and Kandy districts: The first detailed investigation in Sri Lanka. Res Sq. 2020.

23. Moreno-López S, Pérez-Herrera LC, Peñaranda A. Designing a multidimensional vulnerability index for supervising dengue cases from 2015 to 2020 in a low/middle-income country: A spatial principal component analysis. PLoS Negl Trop Dis. 2025;19: e0013556. doi:10.1371/journal.pntd.0013556

24. Instituto Nacional de Salud. Portal Sivigila 4.0. 2024 [cited 4 Aug 2025]. Available: https://portalsivigila.ins.gov.co/

25. Instituto Nacional de Salud. Protocolo de Vigilancia en Salud Pública: Dengue. Bogotá; 2014. Available: https://www.minsalud.gov.co/Documents/Salud%20P%C3%BAblica/Ola%20invernal/Protocolo%20Vigilancia.pdf

26. Departamento Nacional de Planeación. TerriData. [cited 4 Aug 2025]. Available: https://terridata.dnp.gov.co/

27. DANE. Comunicado de prensa. Bogotá; 2025.

28. Departamento Administrativo Nacional de Estadistica. DANE. 2005 [cited 15 Feb 2024]. Available: https://www.dane.gov.co/

29. Instituto Nacional de Salud (Colombia). Portal Sivigila. 2023 [cited 16 Feb 2024]. Available: https://portalsivigila.ins.gov.co/

30. De La Hoz F, Enrique M, Duran M, Vigilancia D, Del Riesgo En A, Pública S, et al. Protocolo de Vigilancia en Salud Pública: Dengue. Bogotá; 2014.

31. Departamento Administrativo Nacional de Estadistica. DANE. 2005 [cited 16 Feb 2024]. Available: https://www.dane.gov.co/

32. TerriData DNP. TerriData DNP. In: TerriData DNP [Internet]. 2023 [cited 16 Feb 2024]. Available: https://terridata.dnp.gov.co/

33. 33. Banco de la República. Banco de la República. In: banrep.gov.co [Internet]. 2022 [cited 15 Feb 2024]. Available: https://www.banrep.gov.co/es

34. 34. Departamento Administrativo Nacional de Estadistica. Geoportal DANE. In: Bogotá [Internet]. 2020 [cited 16 Feb 2024]. Available: https://geoportal.dane.gov.co/#gsc.tab=0

35. 35. World Bank Group. Data Catalog Climate Change Knowledge Portal. 8 May 2024 [cited 7 May 2024]. Available: https://climateknowledgeportal.worldbank.org/download-data

36. 36. NOAA. Climate Prediction Center. In: National Weather Service [Internet]. 2018 [cited 15 Feb 2024] p. 2. Available: https://origin.cpc.ncep.noaa.gov/products/analysis_monitoring/ensostuff/ONI_v5.php

37. Hartig F. DHARMa: Residual Diagnostics for Hierarchical (Multi-Level / Mixed) Regression Models. In: CRAN: Contributed Packages [Internet]. Comprehensive R Archive Network (CRAN); 18 Oct 2024 [cited 11 Sep 2025]. doi:10.32614/CRAN.PACKAGE.DHARMA

38. 38. CRAN: Package mgcv. [cited 5 Aug 2025]. Available: https://cran.r-project.org/web/packages/mgcv/index.html

39. Wood SN. Generalized Additive Models: An introduction with R. 2nd ed. Press C, editor. Text in statistical science. Boca Raton; 2017.

40. Whiteman A, Mejia A, Hernandez I, Loaiza JR. Socioeconomic and demographic predictors of resident knowledge, attitude, and practice regarding arthropod-borne viruses in Panama. BMC Public Health. 2018;18: 1–13. doi:10.1186/s12889-018-6172-4

41. Ebi KL, Nealon J. Dengue in a changing climate. Environ Res. 2016;151: 115–123. doi:10.1016/j.envres.2016.07.026

42. Mulligan K, Dixon J, Joanna Sinn C-L, Elliott SJ. Is dengue a disease of poverty? A systematic review. Pathog Glob Health. 2015;109: 10–18. doi:10.1179/2047773214Y.0000000168

43. Bavia L, Melanda FN, de Arruda TB, Mosimann ALP, Silveira GF, Aoki MN, et al. Epidemiological study on dengue in southern Brazil under the perspective of climate and poverty. Sci Rep. 2020;10: 2127. doi:10.1038/s41598-020-58542-1

44. Qi X, Wang Y, Li Y, Meng Y, Chen Q, Ma J, et al. The Effects of Socioeconomic and Environmental Factors on the Incidence of Dengue Fever in the Pearl River Delta, China, 2013. PLoS Negl Trop Dis. 2015;9: e0004159. doi:10.1371/journal.pntd.0004159

45. Morin CW, Comrie AC EKC. Climate and dengue transmission: evidence and implications. Environ Health Perspective. 2013;121: 1264–1272.

46. Colón-González FJ, Fezzi C, Lake IR, Hunter PR. The Effects of Weather and Climate Change on Dengue. PLoS Negl Trop Dis. 2013;7. doi:10.1371/journal.pntd.0002503

47. Mordecai EA, Cohen JM, Evans M V., Gudapati P, Johnson LR, Lippi CA, et al. Detecting the impact of temperature on transmission of Zika, dengue, and chikungunya using mechanistic models. PLoS Negl Trop Dis. 2017;11: 1–18. doi:10.1371/journal.pntd.0005568

48. Karim MN, Munshi SU, Anwar N, Alam MS. Climatic factors influencing dengue cases in Dhaka city: A model for dengue prediction. Indian J Med Res. 2012;136: 32. doi:10.1016/j.ijid.2018.04.3862

49. Udayanga L, Gunathilaka N, Iqbal MCM, Abeyewickreme W. Climate change induced vulnerability and adaption for dengue incidence in Colombo and Kandy districts: the detailed investigation in Sri Lanka. Infect Dis Poverty. 2020;9: 102. doi:10.1186/s40249-020-00717-z

50. Wu X, Lu Y, Zhou S, Chen L, Xu B. Impact of climate change on human infectious diseases: Empirical evidence and human adaptation. Environ Int. 2016;86: 14–23. doi:10.1016/j.envint.2015.09.007

51. Márquez Benítez Y, Cortés Monroy JK, Martínez Montenegro EG, Peña-García VH, Monroy Díaz ÁL. Influencia de la temperatura ambiental en el mosquito Aedes spp y la transmisión del virus del dengue. CES med. 2019;33: 42–50.

52. Gomes AF, Nobre AA, Cruz OG. Temporal analysis of the relationship between dengue and meteorological variables in the city of Rio de Janeiro, Brazil, 2001-2009. Cad Saude Publica. 2012;28: 2189–2197. doi:10.1590/s0102-311x2012001100018

53. Gómez Gómez RE, Kim J, Hong K, Jang JY, Kisiju T, Kim S, et al. Association between Climate Factors and Dengue Fever in Asuncion, Paraguay: A Generalized Additive Model. Int J Environ Res Public Health. 2022;19. doi:10.3390/ijerph191912192

54. Watts MJ, Kotsila P, Mortyn PG, Sarto i Monteys V, Urzi Brancati C. Influence of socio-economic, demographic and climate factors on the regional distribution of dengue in the United States and Mexico. Int J Health Geogr. 2020;19. doi:10.1186/s12942-020-00241-1

55. Nguyen LT, Le HX, Nguyen DT, Ho HQ, Chuang TW. Impact of climate variability and abundance of mosquitoes on dengue transmission in central Vietnam. Int J Environ Res Public Health. 2020;17. doi:10.3390/ijerph17072453

56. Schmidt W-P, Suzuki M, Dinh Thiem V, White RG, Tsuzuki A, Yoshida L-M, et al. Population Density, Water Supply, and the Risk of Dengue Fever in Vietnam: Cohort Study and Spatial Analysis. Farrar J, editor. PLoS Med. 2011;8: e1001082. doi:10.1371/journal.pmed.1001082

57. Naish S, Dale P, Mackenzie JS, McBride J, Mengersen K, Tong S. Climate change and dengue: A critical and systematic review of quantitative modelling approaches. BMC Infect Dis. 2014;14: 167. doi:10.1186/1471-2334-14-167

58. Lippi CA, Stewart-Ibarra AM, Muñoz ÁG, Borbor-Cordova MJ, Mejía R, Rivero K, et al. The social and spatial ecology of dengue presence and burden during an outbreak in Guayaquil, Ecuador, 2012. Int J Environ Res Public Health. 2018;15. doi:10.3390/ijerph15040827

59. Udayanga L, Gunathilaka N, Iqbal MCM, Lakmal K, Amarasinghe US, Abeyewickreme W. Comprehensive evaluation of demographic, socio-economic and other associated risk factors affecting the occurrence of dengue incidence among Colombo and Kandy Districts of Sri Lanka: A cross-sectional study. Parasit Vectors. 2018;11: 1–18. doi:10.1186/s13071-018-3060-9

60. Atique S, Abdul SS, Hsu CY, Chuang TW. Meteorological influences on dengue transmission in Pakistan. Asian Pac J Trop Med. 2016;9: 954–961. doi:10.1016/j.apjtm.2016.07.033

61. Huynh LTM, Stringer LC. Multi-scale assessment of social vulnerability to climate change: An empirical study in coastal Vietnam. Clim Risk Manag. 2018;20: 165–180. doi:10.1016/j.crm.2018.02.003

62. Escobar-Mesa J, Gómez-Dantés H. Determinantes de la transmisión de dengue en Veracruz: Un abordaje ecológico para su control. Salud Publica Mex. 2003;45: 43–53. doi:10.1590/S0036-36342003000100006

63. Moreno-López S, Pérez-Herrera LC, Peñaranda A. Designing a multidimensional vulnerability index for supervising dengue cases from 2015 to 2020 in a low/middle-income country: A spatial principal component analysis. Mang’era C, editor. PLoS Negl Trop Dis. 2025;19: e0013556. doi:10.1371/JOURNAL.PNTD.0013556

64. Hales S, de Wet N, Maindonald J, Woodward A. Potential effect of population and climate changes on global distribution of dengue fever: an empirical model. Lancet. 2002;360: 830–834. doi:10.1016/S0140-6736(02)09964-6

65. Tapia-Conyer R, Betancourt-Cravioto M, Mendez-Galvan J. Dengue: an escalating public health problem in Latin America. Paediatr Int Child Health. 2012;32 Suppl 1: 14–17. doi:10.1179/2046904712Z.00000000046

66. Maestre RGD. Dengue: epidemiología, políticas públicas y resistencia de vectores a insecticidas. Rev Cinc Biomed. 2013;4: 302–317.

67. Bardach AE, García-Perdomo HA, Alcaraz A, Tapia López E, Gándara RAR, Ruvinsky S, et al. Interventions for the control of Aedes aegypti in Latin America and the Caribbean: systematic review and meta-analysis. Tropical Medicine and International Health. 2019. doi:10.1111/tmi.13217

68. Lee SA, Jarvis CI, Edmunds WJ, Economou T, Lowe R. Spatial connectivity in mosquito-borne disease models: a systematic review of methods and assumptions. J R Soc Interface. 2021;18. doi:10.1098/RSIF.2021.0096

69. World Health Organization. Dengue: Guidelines for Diagnosis, Treatment, Prevention and Control. Genova; 2009.

